# Adapting to scarcity: plasticity in rural healthcare practice

**DOI:** 10.64898/2026.03.14.26348407

**Authors:** Crystal Gaudet, Don Eby, Lisa Shepherd, Frances Kilbertus, Erin Kennedy, Sayra Cristancho

## Abstract

Rural healthcare practitioners face persistent challenges such as limited staffing, geographic isolation, and broad scopes of responsibility. These conditions require constant adaptation to ensure care delivery. This study introduces the concept of plasticity, adapted from sociobiology; to examine how rural healthcare teams collectively navigate scarcity through flexible role and task shifting. Drawing on constructivist grounded theory, we conducted 19 interviews with nurses and physicians in emergency departments in two rural communities in Ontario, Canada, to explore how plasticity functions in rural healthcare teams. The analysis identified two variations of plasticity: acute (short-term, high-stakes task switching or role expansion) and chronic (long-term role expansion), alongside four interrelated dimensions that characterize this phenomenon in rural settings. While acute plasticity was often empowering, chronic plasticity, exacerbated by the COVID-19 pandemic, contributed to cumulative stress, burnout, and professional demoralization. Our analysis illustrates that plasticity is both a strength and vulnerability in rural healthcare work, sustained through informal learning and relational responsibility to community, yet largely unsupported by formal institutional structures.

## 1. Introduction

Rural healthcare is complex, shaped by factors such as low population density, geographic isolation, limited resources, broad scopes of practice and responsibilities, harsh weather, and the overlap of personal and professional relationships (Kolhatkar et al., 2017; MacKinnon, 2012; Strasser, 2016). These conditions require health care practitioners (HCPs) in rural areas to adapt to effectively meet the needs of their communities (Warner et al., 2005). One key adaptation identified in the literature is the adoption of fluid roles and flexible responsibilities, driven by reduced human and physical resources (Pesut et al., 2014; Scharff, 2013). As a result, rural HCPs move away from rigid roles and embrace a generalist approach, understood as a “commitment to a breadth of practice, within teams, to meet the needs of patients and communities” (Kelly et al., 2021, p. 786). This often involves taking on tasks their urban counterparts might not typically perform (Chipp et al., 2011; Kolhatkar et al., 2017; Pesut et al., 2014; Strasser, 2016; Whiteing et al., 2022), reconfiguring responsibilities within rural healthcare teams to ensure essential services are maintained despite resource constraints.

In rural healthcare, both nurses and physicians frequently adjust their roles, assuming additional duties or temporarily switching tasks to maintain care delivery (Scharff, 2013; Setrinen Hansen et al., 2021). For example, nurses may initiate emergency protocols until a physician arrives (Scharff, 2013), while physicians might take on tasks typically performed by nurses, such as patient coordination, to maintain workflow (Setrinen Hansen et al., 2021). Weather disruptions and transport delays further reinforce the need for such flexibility. Beyond swapping tasks, rural HCPs often assume multiple roles to fill gaps in care and meet community specific needs (Chipp et al., 2011; Scharff, 2013; Walkerman, 2008). In the absence of specialists, for instance, rural physicians may perform procedures outside their formal training (Falk et al., 2020; Konkin et al., 2020), while nurses develop expertise across multiple specialty areas (MacKinnon, 2012; Scharff, 2013). Task shifting from physicians and allied health professionals to nurses has also been a longstanding strategy for addressing staff shortages in rural contexts (Feringa et al., 2018; Martin-Misener et al, 2020; McCallum et al., 2024; McCullough, 2022). Consequently, the generalist orientation of rural practice requires HCPs to embrace flexibility and versatility, both individually and collectively, to sustain care amid evolving demands.

Despite their prevalence, these adaptations are often framed as individual or professional resilience, obscuring how scarcity is structurally produced and how its burdens are redistributed onto providers. A conceptual lens is therefore needed not only to name these practices, but to clarify what they reveal about system accountability and the organization of healthcare work under conditions of constraint. Although flexible and expanded roles are well documented in rural healthcare, they are frequently examined in isolation, limiting insight into their collective dynamics and cumulative effects. Looking beyond healthcare, the concept of plasticity from sociobiology offers a useful a lens for understanding how rural healthcare teams collectively adapt to scarcity.

Plasticity describes how superorganisms, like social insects, collectively respond to disruption by adapting or interchanging roles (Cristancho and Thompson, 2023). In stable environments, social insects like bees and ants maintain a strict division of labor, with individuals assigned to specialized roles. However, in the face of disruption, roles become fluid. For example, in honeybee colonies, worker bees, typically non-reproductive, can take on egg-laying duties if the queen is lost. Likewise, nurse bees can step in as foragers or guards when needed (Cristancho and Thompson, 2023). As Cristancho and Thompson note, these collective responses are driven by a common purpose: “to keep the colony alive and reproducing as long as possible” (p. 254).

In this sense, plasticity is not merely individual flexibility, but a collective behaviour oriented toward sustaining a shared purpose. Building on this work, we suggest that plasticity offers a powerful metaphor for understanding how rural healthcare teams collectively sustain generalist practice in the face of constrained resources and shifting demands.

Previous research has applied plasticity to examine fluid teams across domains, highlighting its role in supporting cohesion, problem solving, and adaptability, while also raising concerns about unintended risks (Cristancho et al., 2022). Although this work focused primarily on urban healthcare teams, rural contexts were identified as settings in which plasticity may be especially pronounced, underscoring the need for further exploration.

The unique demands of rural practice make it an ideal setting to explore the full scope of plasticity: its advantages and its risks. This study therefore examines how plasticity shapes the work of rural healthcare teams, offering a language to articulate collective adaptation under conditions of scarcity. Clarifying plasticity contributes to a deeper understanding of how generalist rural healthcare work is organized and sustained.

## 2. Methods

### Defining Rurality

Defining rurality presents ongoing challenges and can vary with perspective (Simpson and McDonald, 2017). Common definitions emphasize factors such as population size or density, geographic distance from urban centres, and reliance on informal or community-based services. For this study, we conceptualize rural health care as a distinct practice context characterized by specific structural and relational features (Fors, 2023; Strasser, 2016). Rural health care is generalist in scope, grounded in primary care, and constrained by limited access to specialist services and advanced technologies. Compared to urban counterparts, rural health care practitioners often assume broader clinical responsibilities, work with fewer supports, and experience greater professional isolation (Strasser, 2016). They are also more likely to be embedded within the communities they serve, navigating multiple social roles and overlapping personal and professional relationships with patients and families, in contrast to urban settings where care is more often delivered by relative strangers (Fors, 2023; Simpson and McDonald, 2017).

### Setting

This study was conducted in two rural communities in Ontario, Canada, each with a Rurality Index for Ontario (RIO) score greater than 65. The RIO is a measure of rurality based on population size, population density, and travel time to both basic and advanced referral centres. It is used provincially to determine eligibility for certain programs and incentives tied to degree of rurality. Scores range from 0 (most urban) to 100 (most rural), with scores above 40 often qualifying for various supports. Both study sites exceed this threshold. Emergency departments in both communities operate with single-physician coverage, supported by nurse practitioners (NPs), nurses (RNs), or registered practical nurses (RPNs, known elsewhere as Licensed Practical Nurses).

### Study Design

Guided by constructivist grounded theory (CGT), this study aimed to develop a conceptual understanding of how plasticity unfolds within rural health care teams. Plasticity was treated as a sensitizing concept to explore how teams collectively adapt roles and responsibilities in response to disruption and constraint. CGT provided the methodological approach for examining how this social process is enacted in practice, particularly within rural settings where structural limitations, workforce composition, and community-embedded practice shape everyday care delivery.

Semi-structured interviews explored team composition, the types of care provided, and experiences of adapting roles and responsibilities in rural emergency settings. Participants were introduced to a provisional working definition of plasticity and invited to reflect on its usefulness, desirability, and limits in describing their team practices. They were also asked to describe concrete situations in which roles shifted, and the factors that facilitated or constrained these adaptations (see Supplemental Material).

### Participants and Recruitment

Participants were recruited through purposive and snowball sampling methods (Kuzel, 1992), guided by theoretical sampling. Based on prior research suggesting that rural emergency settings are particularly generative sites for examining plasticity, recruitment focused on emergency departments in the two study communities. Between January and November 2024, a total of 19 semi-structured interviews were conducted with 9 nurses (6 RNs, 2 RPNs, 1 NP) and 10 physicians, two of whom also held administrative roles. Participants’ clinical experience ranged from fewer than 5 years to over 40 years. To protect confidentiality, participants are referred to by study IDs (e.g., N3, P6), where “N” denotes nurse participants and “P” denotes physician participants.

### Data Collection

Interviews lasted 60–90 minutes and were conducted virtually via Zoom by three members of the research team [initials omitted for double-anonymized peer review]. Interviews were audio-recorded and transcribed verbatim. All participants provided written informed consent. Ethics approval was granted by the institutional review board of the researchers’ affiliated institution [details omitted for double-anonymized peer review].

### Data Analysis

Data were analyzed using the constant comparative method (Charmaz, 2014). Three members of the research team independently conducted line-by-line coding of the first three transcripts, then met to develop an initial coding framework. This framework was applied to subsequent transcripts using Quirkos software, with categories iteratively refined, merged, or expanded as analysis progressed.

Consistent with CGT, data collection and analysis were conducted concurrently and were guided by theoretical sampling. The research team met after every five to six interviews to compare interpretations and identify areas requiring further elaboration, which informed both recruitment and interview questions (e.g., recruiting participants with administrative responsibilities and attending more closely to informal learning practices). A return-of-findings process was used to explore resonance between emerging interpretations and participants’ experiences. Data collection continued to theoretical sufficiency, when categories were well developed and further interviews no longer generated new analytic insights (Charmaz, 2014). The final analysis identified two variations of plasticity (acute and chronic) and four interrelated dimensions that together constitute a grounded theoretical account of plasticity in rural emergency care.

### Research Team and Reflexivity

The research team consisted of six members: two qualitative research experts [initials omitted] and four healthcare practitioners [initials omitted], each bringing diverse perspectives to the study. Three members of the research team [initials omitted] are physicians with experience in both rural and urban settings, and one [initials omitted] is a nurse practitioner with urban experience. This combination of insider knowledge, comparative insights, and outsider perspectives enriched both the study design and its interpretation.

## 3. Results

The analysis of participant interviews identified two variations of plasticity, acute and chronic, and four interrelated dimensions that characterize how plasticity is enacted within rural emergency departments. These variations and dimensions capture the structural and relational conditions that shape team-level adaptation, providing a framework for understanding the phenomenon. The following results section illustrates this framework through participants’ accounts, showing how plasticity operates as a collective adaptive capacity in under-resourced rural emergency care.

### 3.1 Acute and Chronic Plasticity

Plasticity shaped the daily dynamics of interprofessional work in the rural emergency departments examined in this study. Teams were typically composed of one physician and one or two nurses, though their configuration shifted constantly according to patient needs, transfers, travel, and time of day. At times, one nurse might be staffing the emergency department, another accompanying a patient by ambulance to a larger centre, or the physician might be on call at home throughout the night. Within this context, participants emphasized the importance of being “prepared to shift roles and share power and move into each other’s kind of tasks” (P9).

Plasticity was described as a normalized feature of rural practice. As one physician explained:

> *“So, if that nurse is like doing this point of care lab testing, but if I can get the other patient […] a Gravol […] and at least start their treatment, then I’ll do that, right? So, I think we have a lot of role flexibility like sideways and around that maybe isn’t seen in larger departments” (P1).*

For participants, enacting plasticity meant “do[ing] what needs to be done” (P8) to keep the emergency department functioning. This could involve interchanging roles, expanding one’s responsibilities, or layering additional tasks in response to environmental or systemic disruption.

For example, a physician might clean the room after a resuscitation because no cleaning staff are available at night, or, as another physician recalled:

> *“I’ve gowned up to wipe poop off [a] patient […] to help the nurse because there’s only me and a nurse and if there’s nobody else in the department, then like I’m not above […] helping out to make the flow better, right?” (P1)*

Enacting plasticity, then, meant stepping outside one’s usual role, temporarily interchanging roles with colleagues, or absorbing tasks not otherwise one’s responsibility to sustain team functioning. Importantly, these actions were consistently framed as team-based efforts. As one nurse explained to a student who questioned whether feeding patients was part of the RN’s role:

> *“We’re feeding the patients down the hallway because we’re a team. We got to work as a team, right? That’s what we do” (N5).*

Our initial conceptualization of plasticity focused on acute plasticity: voluntary, unconventional, heroic actions in response to situations requiring timely intervention. In this study, acute plasticity most often occurred during episodic, urgent or time-sensitive situations when the provider was required to step outside of their comfort zone. Physicians’ stories of *“being family physicians doing specialist work”* (P7) or nurses’ stories of *“x-ray techs jump[ing] in on codes with us”* or police offering *“an extra set of hands even if it’s just taking notes for us”* (N5) exemplified acute plasticity. Such instances required the willingness and courage to take on tasks beyond what would normally fall within their broad scope of practice, including procedures they had little experience with, or had only observed, but were compelled to attempt when the risk of harm was imminent if no one acted. Yet, participants noted that such acute situations were relatively rare. As one physician observed:

> *“we can talk about like the trauma patient that comes in and you’re uncomfortable with and they need all these interventions. It’s relatively uncommon, thankfully, but what ends up being common is we end up being the social worker. We end up being the distribution manager” (P1).*

Their accounts highlighted a more sustained, non-voluntary variation: chronic plasticity. Unlike acute plasticity, which was temporary and situational, chronic plasticity referred to the ongoing expectation, especially for nurses, to perform multiple, non-traditional tasks as a routine feature of their work due to limited staffing. These tasks included clerical work, laboratory testing, mixing medications, housekeeping, and even cooking for patients, as one nurse described:

> *“We answer phones. We make appointments for people that are calling in for their blood work. We assist with casting. I’m dealing with public health because something’s gone wrong with our immunization fridge. We don’t have maintenance on site. So, I’m dealing with heat issues back in x-ray. We don’t have a ward clerk. So, I’m processing doctor’s orders, filling out forms, getting people prepped for CTs, arranging transport, drawing blood because we don’t have lab as well”* (N4)

With fewer human resources compared to larger centres, nurses described performing multiple roles simultaneously and needing to be skilled in several different areas. This added extra layers to the nurses’ routine tasks, such as *“hav[ing] to take on the administrative role at the same time as you’re also trying to care for the patient”* (N1). As one nurse shared, the chronicity of plasticity in rural emergency departments was well depicted by a local cartoonist (Figure 1).

**Figure 1:**
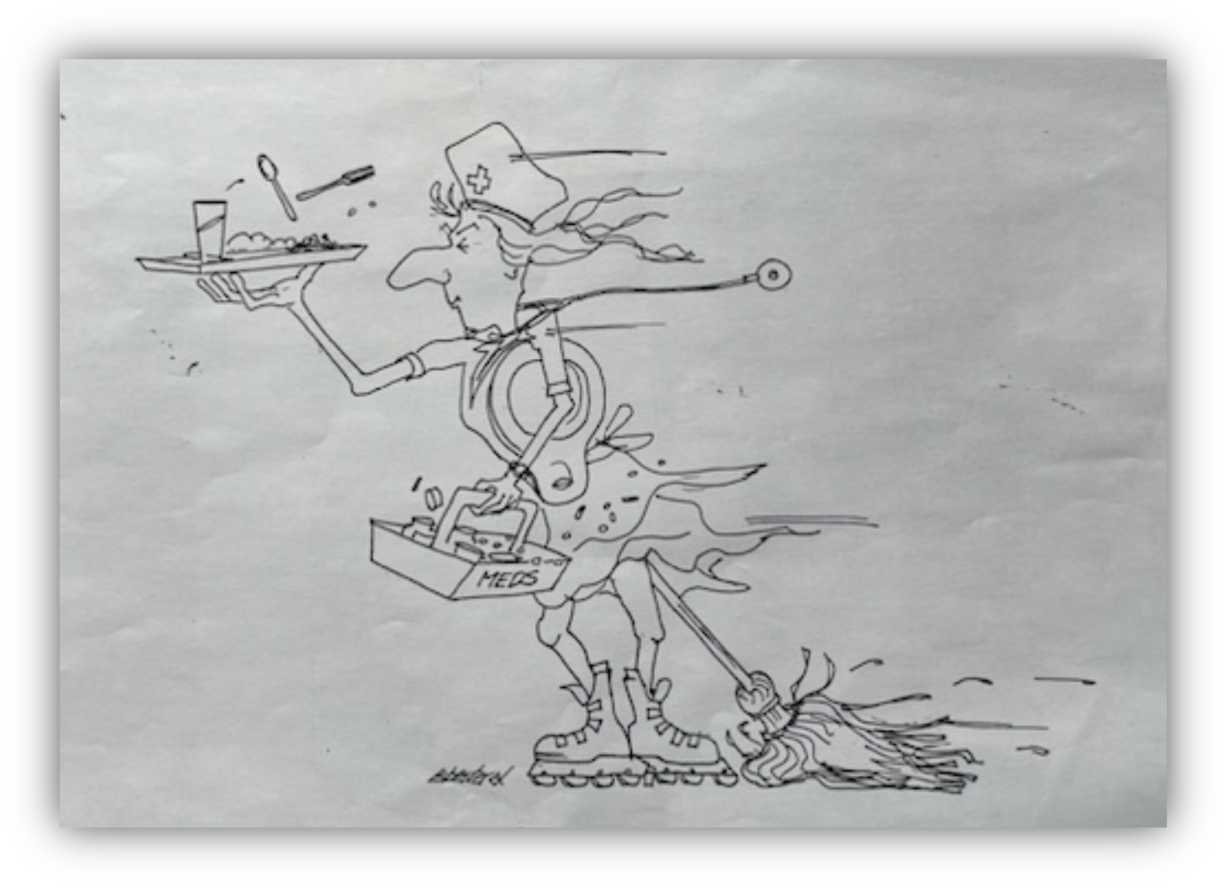
Cartoon created by a local community member depicting the diverse tasks performed by nurses in their community, illustrating the chronicity of plasticity in rural emergency departments.

This chronic performance of non-traditional tasks that competed with and sometimes jeopardized nursing tasks, was highly stressful and demoralizing because, as one nurse explained, *“you’ve got all these demands and you’re trying to do all of this stuff, so you just feel it pushing you down and pushing you down”* (N2). Despite the pressure, most nurses indicated that *“it’s nice to be able to help people […] no matter what the problem is as a nurse”* (N7). Thus, plasticity in rural practice was referred to by both nurses and physicians as a day-to-day struggle and reward.

Together, acute and chronic plasticity demonstrated how rural emergency teams adapted to sustain service delivery and team functioning in under-resourced settings, while also exposing the limits of these practices. The next section outlines four dimensions that further characterized how plasticity was enacted in rural environments.

### 3.2 Rural plasticity challenges assumptions around “best care”

The variations of plasticity described above not only illustrate how rural teams adapted but also demonstrated how these adaptations challenged conventional assumptions about what constituted “best care.” The differences between rural and urban practices were a point of contention for most participants, who felt that norms and standards established in urban-based practice did not reflect the realities of rural practice. Therefore, through their accounts of plasticity, participants emphasized the message that rural care *“isn’t poor care; it is the best care they’re going to get where they are”* (P8). One physician described this as the impossibility of *“comparing what’s happening with an ideal that is not on the table”* (P3).

Several physicians elaborated on this tension through detailed accounts of acute situations that required them to try something that in a bigger hospital they would not be required to do because they have access to specialized teams and departments. These stories underscored how plasticity was not about lowering standards but about making ethically necessary choices to ensure patients received timely care. As one physician explained:

> *“So I think that regularly, you are the doc who, […] you’re literally the only one in the hospital, so if you have reasonable confidence in your ability to do something, even if you’ve never done it before, then you do it, because otherwise you’re sending people four hours, six hours, they can wait days to get out of town, if the weather’s bad, forget it, they’re going nowhere, so you just do it unless it seems really reckless or unsafe” (Phy 4).*

Here, plasticity was not understood as a compromise, but as an ethically necessary adaptation that enabled patients to receive care that would otherwise be inaccessible.

Additionally, for most participants, their close involvement with the community and its associated expectations were also part of what they needed to consider to provide care in rural environments. On one hand, healthcare practitioners’ actions are easily known, as illustrated by one participant’s account:

> *“If you live in a small place, you’re not a stranger, your professional life is something that’s known. So, if you mess up badly, everybody will know. […] even if it’s just like, you were a jerk to somebody in the ER, or you were not that sympathetic. That’s gonna be common knowledge in the village”* (P7)

On the other hand, this visibility made many participants aware that *“there are things that you might have to do as a rural doc that you don’t have to do in urban centers”* (P5), based on the expectation of patients who *“just want you to do it”* (P1). And as one nurse reflected, these expectations demonstrated the understanding of the community as to what’s possible or not for their healthcare practitioners:

> *“I think the community also understands that we aren’t always working within the most comfortable, ideal settings. So, I think the expectations from people are realistic for the most part, as far as we’re generalists and we’re sometimes working in areas that aren’t our most favorite or most knowledgeable”* (N8)

Examples like these prompted reflection in many participants who felt that this reality posed a challenge to health care practitioners new to rural practice or practicing as locums or travel nurses, as it differs from what they learned in urban-based educational environments. Several participants described their experiences of working with “*locums that have only ever practiced in urban places and that really struggle with the breadth of what you’re expected to do”* (P5). For instance, as one physician noted, some locums *“would insist on having something done that the nurses just didn’t have time for right at that time. So, they would wait for it to be done rather than doing it themselves”* (P6). Nurses similarly observed that for travel nurses hired through agencies on short-term contracts, *“there’s not the same community type commitment or investment in the community, as far as doing the extras going above and beyond”* (N8). Thus, participants linked their willingness to expand roles to their embeddedness in the community, which fostered a sense of responsibility they felt was often lacking among those working only temporarily in the setting.

Given the reality of rural practice, plasticity was seen by participants as essential to providing the best possible care for their community. Their accounts underscored how plasticity both challenged urban-based assumptions of best care and reflected community-embedded expectations of responsibility and adaptability. Their accounts also highlighted the consequences of plasticity for rural practitioners, including both its rewards and its strains.

### 3.3 Plasticity in rural practice brings rewards, yet takes a toll

In addition to reshaping notions of best care, participant accounts provided insight into the personal impact of plasticity. When plasticity was a voluntary act, meaning participants had the autonomy to adapt their roles based on interest, willingness, or for professional growth, it generally promoted positive feelings among participants. It made them feel exhilarated because, as one physician explained, *“you’ve gone beyond, you’ve done what you had to do, and there’s been a good outcome”* (P8). It kept them engaged, as it allowed them to expand their knowledge and skills, as a nurse reflected: *“you feel a bit like a fish out of water, but I do trust that it’s all going to come together […] I feel like […] it’s just good for my brain to be challenged”* (N7).

Participants also spoke positively about how plasticity prompted a different approach to practice because *“there’s just you and one other person; there is no one else. So, you have to bend”* (N3). This was particularly evident in trauma situations when physicians could be *“putting lines on the patient, getting the blood pressure, trying to start an IV, because my colleagues* [an RN and an RPN] *are also busy doing things”* (P3), which contrasted with their experiences of urban practice with ample resources. As one physician described,

> *“in the boonies, […] the nurse isn’t taking orders from you, because she’s focused on trying to get that IV in and you can say all the orders you want, they’re not going anywhere. You gotta help her do the nursing tasks to get that initial resuscitation going. I quite like it [because] for me at least it helps with the collegiality at my hospital”* (P3).

However, particularly for nurses, plasticity was perceived as an expectation due to limited human resources. For example, two junior nurses might be assigned to the same shift during *“times where there’s gaps to be filled, and anyone they can find to fill them, is better than no one”* (N8). The demands of plasticity became overwhelming and emotionally draining when nurses were torn by needing to be in two places at once, as this participant described:

> *“I’m in emergency helping resuscitate this gentleman. And the phone’s ringing and we can’t answer the phone because we’re all involved in this resuscitative effort. […] when we finally get the ET-tube in we can answer the phone. And it’s my manager. And I’m almost crying on the phone. Cause I said, we need help. We couldn’t get extra help. We couldn’t get another nurse to come in. So, I could have quit that night”* (N2).

This expectation of flexibility required participants to adopt coping strategies to manage stress, including letting things go, setting boundaries, and prioritizing competing demands.

Beyond individual strategies, participants underscored the importance of collegial support. As one nurse reflected:

> *“I like the people that I work with. I think that’s what makes it, even though everyone at times can be really stretched quite thin and people can be stressed, but if we help each other, we’re like, okay, like I need to do this. Can you help me with this? And then I’ll help you do that. We really do work together” (N7).*

For many, peer support was essential in helping them manage multiple demands. Indeed, one of the most common coping mechanisms described across the interviews was having a network of colleagues to whom they could talk and informally learn from. This reliance on peer networks pointed directly to the role of informal learning in enabling plasticity.

### 3.4 Plasticity reveals the pre-eminence of informal training

Coping with the demands of plasticity relied heavily on support from colleagues, and participants consistently emphasized the primacy of informal learning networks in enabling them to adapt. The ability to step into unfamiliar roles was rarely developed through formal training; instead, it was cultivated through peer debriefs, peer mentorship and “just in time” knowledge-sharing. Participants frequently indicated that the formal training systems in place for medicine and nursing were inadequate as preparation to work in rural settings. They were required to do many tasks that were infrequently performed or that fell outside their usual generalist scope of practice. Opportunities for additional training were also limited, since funding and shift coverage were rarely available. In this context, participants relied on one another to develop the skills needed to function effectively in their work environment.

According to most participants, these informal ways of learning were crucial in *“learning how to do the other jobs”* (N4) because for everyone, *“you’re just trying to do the best that you can with what you have”* (N7). The overarching approach was informal rather than formal, as illustrated by one nurse participant:

> *“And when stuff does happen, we will talk about it as a group for the next three or four days of what we did, what we could have done, was the outcome good or not, what could have changed. And then if somebody struggled with the vent[ilator], so then we would informally on nights go over the vent and switch the settings. But it’s all done informally.”* (N6)

Besides teaching and debriefing each other while working together, participants also mentioned the importance of being able to contact peer colleagues, who were not working at the time, to help them when they needed it. For instance, while camping with her family, a nurse taught her colleague, over FaceTime, how to work a rarely used piece of equipment needed to treat a patient. While *“that’s not something you can do every day”* (N1), participants described it as necessary because *“it’s something that is hard to teach outside of experiential learning, so it’s often learning from your colleagues who have done it for more years than you”* (N8).

In addition to peer support, participants emphasized the internet, particularly YouTube, as a practical tool for ‘just in time’ learning of specific skills and techniques. Physicians described turning to online videos to quickly refresh rarely performed or unfamiliar procedures outside their routine generalist practice. As one explained, “*Maybe I hadn’t done some sort of reduction, or a particular procedure and you sort of teach yourself. It’s gotten a lot easier now with YouTube. Before I was pulling out books and doing it”* (P6). Another physician added, *“there’s some great teachers providing instruction on YouTube”* (P8). Such accounts highlighted how, in the absence of readily available specialist training or supervision, practitioners drew on informal online resources to adapt in the moment, maintaining competence and ensuring the patient still received care.

Their accounts also illustrated how participants saw themselves as generalists rather than specialists, emphasizing the need *“to know a lot about a lot of things”* while also acknowledging that *“you can’t know everything”* (N3). One physician elaborated on this challenge reflecting: *“I think that it’s harder to be a generalist in many ways because you have to be comfortable with being uncomfortable. You’re never going to be amazing at everything”* (P1). The qualities required of a rural generalist were described as a key driver in enacting plasticity, providing a foundation for stepping into unfamiliar roles or tasks. However, most participants expressed concern that neither the generalist role nor the informal learning that sustained plasticity were explicitly acknowledged.

### 3.5 Plasticity demands recognition by the system

While informal networks enabled practitioners to enact plasticity, their accounts indicated that relying solely on personal willingness and peer support was unsustainable. Many participants’ willingness to enact plasticity, both in acute and more chronic scenarios, was motivated by a sense of responsibility to provide the best care possible to community members. As one physician explained, this responsibility to the community “*opens up your interest or ability to be able to sort of take on things that don’t feel entirely comfortable*” (P5). However, some expressed concerns that if organizations continue to rely solely on the personal willingness of its members, it risks losing respect and sustainability in the long run. As one physician described it:

> *“If the system works, then […] you’re able to draw on all those resources. And if all those people that you reach out to ask for help from are completely overworked,[…] then I can do very little”* (P4)

During the interviews, the organizations these individuals work for acknowledged the difficulty of working in a *“resource negative”* (P10) environment. For instance, there was an awareness that nurses were doing many different tasks that were not scripted into their job description, which translated into “*worries when the new nurses are coming in here, how are they going to look at these expectations”* (P10).

Many participants emphasized that physician and nurse shortages, especially following the Covid-19 pandemic, created conditions in which organizations are *“working short chronically; we’re having [patient] mental health and addictions chronically; we’re having chronic violence”* (N9). In response, organizations often resorted to hiring locums and travel nurses, which created unintended consequences for existing staff. As one nurse noted, *“we never had travel nurses until three years ago and they don’t embed themselves in the culture or the community. They sit off in the nurse’s station. It’s really impacted the team a great deal”* (N9). Even when organizations attempt to *“grow their own and infuse them into the environment so they become accustomed to the expectations”*, there was recognition that *“I don’t think we have enough local people to really bring back”* (N9).

In this context, participants described a system under strain where both individuals and organizations were affected. Organizations struggled to find the resources to recognize the efforts and alleviate the workload of individuals who go above and beyond to provide care for *“the patient in front of you, the community, trying to make sure that things work out as best as possible”* (P4). Some participants revealed that they felt at the breaking point because of the organization’s struggle with retention. According to some nurses, new people *“don’t have the skillset that our staff, as RPNs have”* (N3) and therefore they leave to work in places where those tasks are not the expectation.

This willingness to help despite being overworked was vividly described by various participants as an *“elastic that’s just stretched way too thin”* (P8). In many instances, individuals found themselves having to *“pick yourself up, give yourself a pep talk and say I can figure this out”* because they were also aware that *“you’re also there to help, right? Obviously if they need the staff, they need the staff and you’re another person whose kind of helping the bigger picture”* (N7). While this organizational issue was very frustrating for many participants, some recognized that their environments were not as busy as larger centres, and they understood the difficulty for the organization to justify putting in place more resources, as this physician illustrated:

> *“We made the case many times that we should have a second nurse in ER at night, but their hands are tied too. They only have so much funding, so I don’t think that it’s lack of support. It’s just, there’s nothing they can do”* (P8).

Besides lack of resources, most participants highlighted the lack of appreciation from the larger system. For instance, interactions with colleagues in larger centers during patient transfers were described as demoralizing and frustrating when those colleagues made uninformed judgements, as one nurse illustrated:

> *“So, we’ll get somebody come in, we’ve stabilized them, we’ve got to take them to [city]. […] And you get to the ICU, and they’re nitpicking on where your lines are, or how you have it programmed in the pump and it’s like, well “that’s not optimal” and “what you would have done”… but we do what we can. It’s not perfect all the time but the patient is alive now. They weren’t an hour and a half ago”* (N6).

These judgements were often attributed to a lack of knowledge or interest in learning about the rural reality. Participants indicated that because of this lack of knowledge, *“they don’t necessarily appreciate that I’m also answering the phone and doing all this stuff, and that’s why I forgot to call you to tell you that the patient left 20 minutes ago”* (N5).

Overall, participants, as individuals who are part of the system, felt as if their plasticity efforts were invisible and expected. As one physician explained, the perception was that “*these other jobs are considered more menial*, *so no one is patting you on the back saying, wow, you really went above and beyond today”* (P8). As such, many voiced their desire that *“it has to be recognized when it’s going on longer than it should. The chronicity has to be recognized”* (P8) and organizations need to do more to acknowledge and address the plasticity required for rural practice.

## 4. Discussion

This study introduces plasticity as a conceptual lens for understanding how generalist rural healthcare teams adapt under conditions of scarcity. Plasticity frames adaptation not as an individual attribute, but as a collective capacity that sustains team functioning. While not unique to rural healthcare, features of rural practice, including resource constraints, geographic isolation, a generalist orientation and community embeddedness, make plasticity both more prevalent and more visible. Two variations were identified: acute plasticity, involving short-term task interchange or role expansion in urgent situations, and chronic plasticity, the sustained expectation that practitioners, especially nurses, assume non-traditional tasks such as clerical work, laboratory testing, and housekeeping. Acute plasticity was often experienced as rewarding, but chronic plasticity revealed the limits of adaptation and the strain of persistent workforce shortages.

Participants’ accounts of plasticity were informed by a deep sense of responsibility to patients and the communities in which they lived. Their commitment to “doing what needs to be done” reflects an ethical sensibility underpinning their adaptations, consistent with what Fors (2020) calls potato ethics: a moral imperative to be useful, cooperating in whatever way is necessary and learning whatever is required to provide care. Drawing on Tronto’s (2012) theory of relational responsibility, Fors emphasizes that responsibility arises not only from relationships fostered through presence in rural communities, but also from an acute awareness of the rural context and the realities that shape it. In such settings, where providers live and work alongside patients, these responsibilities are intensified. As Fors notes, presence in rural practice entails “sharing the same space and encountering suffering without the distance afforded by bureaucratic structures or narrowly defined professional boundaries” (p. 268). This awareness, Fors argues, drives rural practitioners to be, like the potato, “almost endlessly adaptable” (p. 275). Participants in this study described being constantly aware of their own limitations yet equally conscious of the consequences of failing to act, knowing that without taking on unfamiliar or additional tasks, patients might simply go without care. Plasticity, therefore, was not only a pragmatic response to scarcity but also an ethical practice, grounded in community embeddedness and a relational ethic of care. This moral imperative resonates with sociological analyses of care-oriented communities of practice, which similarly describe repertoires of care driven by a commitment to do “whatever it takes” (Young et al. 2018), underscoring the central role of moral responsibility in sustaining healthcare work under conditions of constraint.

Importantly, many of the tasks participants absorbed, such as laboratory testing, are not included in the legislatively defined nursing scope of practice but are tasks that could technically be performed by anyone. In most nursing practice contexts, however, RNs would not typically take on these responsibilities. What marked them as plasticity was their structural embedding as additional or layered work, assumed because clerks, lab staff, or other supports were unavailable. In this way, chronic plasticity reflects how generalist roles are stretched and reshaped to sustain care under persistent shortages. This builds on MacLeod et al.’s (2019) findings that most rural and remote nurses report working within scope, by showing how systemic shortages normalize “extra” tasks and institutionalize them as chronic expectations.

At the same time, chronic plasticity highlights how such efforts remain largely invisible to the system. Hanlon et al. (2011) describe stealth voluntarism, the unpaid, unrecognized extra work of practitioners in underserviced areas, while Allen (2015) conceptualizes the invisible organizing labour of nurses that sustains healthcare systems but is undervalued. Our findings illustrate how chronic plasticity extends this invisible work, as participants routinely took on clerical, administrative, and technical duties essential to service delivery yet absent from formal recognition. This invisibility reinforces a cycle in which plasticity is assumed rather than supported, normalizing expectations that rural practitioners will continue to “do what needs to be done” without organizational accountability.

Participants further emphasized how the COVID-19 pandemic intensified the chronicity of plasticity, accelerating existing workforce shortages and worsening mental health and addictions crises (Anaraki et al., 2022; Kiran et al., 2022; Rural Ontario Municipal Association, 2024). These pressures are felt most acutely in rural communities, where access to primary care is increasingly limited and emergency departments carry disproportionate burdens. Participants described being stretched beyond their limits, while organizations relied heavily on locums and travel nurses who often struggled to adjust to the plasticity required in rural practice. Rather than relieving pressure, these measures sometimes created further strain for permanent staff.

Coping with these demands depended heavily on informal learning networks. Participants described relying on peer mentorship, collegial support, and just-in-time strategies, including contacting colleagues outside of work or using online resources, to step into unfamiliar roles. While this resourcefulness enabled adaptation, it also exposed gaps in formal training and limited access to continuing education. Consistent with prior research (Kidd et al., 2012; Scharff, 2013; Whiteing et al., 2022), on-the-job learning and peer support emerged as the dominant modes of professional development, yet remained largely unrecognised by formal training systems, and to a lesser extent, hospital organizations.

The dual nature of plasticity, as both rewarding and burdensome, was evident throughout. Acute plasticity could boost morale and collegiality, consistent with prior studies showing that rural HCPs value variety and role expansion (Kolhatkar et al., 2017; Setrinen Hansen et al., 2021). Yet the chronic expectation that practitioners absorb multiple non-traditional roles reflects a systemic failure to address persistent shortages. While earlier research has noted that rural HCPs are often pushed beyond their comfort or competence, it has not problematized these pressures as structural deficiencies (Kidd et al., 2012; Konkin et al., 2020; Whiteing et al., 2022). Taken together, our findings illustrate plasticity not as an individual attribute, but as a collective response enacted within systemic constraints: one that sustains rural care in the face of scarcity while simultaneously exposing the structural neglect of rural health systems.

The findings of this study therefore expose the pitfall of viewing plasticity merely as an individual adaptation: it obscures its systemic roots and masks the deeper structural failures of rural health care. Potato ethics and relational responsibility help explain why rural practitioners enact plasticity, illuminating the moral sensibility that compels them to act in the face of scarcity. At the same time, concepts of invisible labour and stealth voluntarism show how this work is obscured, undervalued, and exploited within health systems. From this perspective, chronic plasticity is less a marker of resilience than a symptom of long-standing underfunding and neglect (Martin-Misner et al., 2020; McCallum et al., 2024). In the absence of institutional recognition, targeted resource allocation, and sustained investment in mentorship and training, continued reliance on practitioner’s willingness to adapt risks positioning plasticity as a mechanism of attrition, rather than a source of resilience.

## Limitations

The theoretical insights developed here may be transferable to other rural health care contexts that share similar conditions of practice, such as small populations, limited resources, and distance from referral centres, and may also hold relevance for larger hospitals facing persistent resource constraints, though always in ways shaped by local context. At the same time, the study is situated specifically in rural emergency departments in Ontario, which may limit transferability to other health care settings or international contexts. Future research could extend this work by examining how plasticity is enacted across a wider range of rural contexts, exploring whether similar forms of adaptation emerge in larger centres under conditions of strain, and investigating how expectations of plasticity are taken up, normalized, contested, or left unexamined across health professions education, organizational socialization, and workforce pathways.

## Conclusion

This study illustrates that while plasticity enables rural health care teams to sustain care under conditions of scarcity, it is not a substitute for structural investment. Acute plasticity demonstrates the commitment and ingenuity of practitioners, but chronic plasticity reveals the hidden costs of relying on invisible labour and relational responsibility to maintain services. In this sense, plasticity is both an ethical practice, rooted in practitioners’ commitment to “do what needs to be done,” and an expression of structural neglect, where health systems depend on adaptation rather than provide adequate support. Plasticity surfaces the social organization of scarcity in rural healthcare - how care is kept going, by whom, and at what cost - shifting attention from individual adaptability to structural accountability. Attending to plasticity as a structural phenomenon is therefore critical not only for understanding recruitment and retention challenges but also the conditions under which rural health systems are sustained over time.

## Data Availability

The data that support the findings of this study are not publicly available due to the potential risk of participant identification within two rural communities.

