## Supplementary material for "Adapting to scarcity: plasticity in rural healthcare practice": Supplmental Material

**Adapting to Scaricty: Plasticity in Rural Healthcare Practice**
**Interview Guide**

1. Can you describe what you do, the composition of the team you usually work with and the type of care you provide?
   - How many years have you been a physician or nurse?
   - What kinds of situations do you face?
   - What kinds of tasks happen?

2. Can you think of a situation you experienced recently where plasticity was enacted? By plasticity we mean taking on other people's tasks or roles that are outside of your normal scope of responsibility, when someone becomes unavailable, or even providing care in an unfamiliar context or a different setting than the traditional one (akin to redeployment during COVID). If you can recall a specific situation where you noticed that the team had to switch roles or tasks or had to perform in an unfamiliar environment to get things done, I am going to ask you to describe it as best you can.

   a. Reflecting on this situation, how do you conceive of the role of plasticity in your team? Is it helpful as a concept for you to understand how your team or group works? Is it desirable why or why not? Would you describe plasticity in another way?
   a. What is required for someone to be able to embrace plasticity in a rural or small community context?
   b. What role does trust play in the practice of plasticity? (Probe for interpersonal and organizational trust if not mentioned)
   c. Have you seen a situation in which plasticity was not being properly practiced or where it was harmful?
   d. Are there organizational policies and procedures that promote or inhibit plasticity in your organization? How do you feel about the support you get from your organization, as it relates to plasticity in clinical practice?

3. In a scenario where you know what needs to be done, you have the skill to do it, it is within your scope of practice, but only under certain conditions. The person who normally does it isn’t there, would you do it? In other words, what is the limit to what you would do? What keeps you from doing it? Can you think of an example?

4. What were your expectations of what the job would be when you started and how does it compare to what you do now?
   a. How do you feel about it?
   b. If participant had training in a rural setting – what drew you to that?
   c. How did you emotionally manage that challenge of being hired for some things but being asked to do more?
   d. What did you do to be able to adapt to the additional expectations of the role?
   e. What can the organization do to help people adapt?

5. In healthcare, “accountability” is a very sensitive word/idea for many. What are your thoughts of the role of “accountability” in allowing or preventing providers to be plastic?

6. We heard from other participants that practicing in a rural or small community context requires you to be a “jack of all trades,” kind of being an expert as a generalist. Do you agree and see yourself as such? If so, how? Based on your experiences, what’s your take on the generalist vs. specialist dilemma?

7. We also heard about people teaching skills to each other. How much of your learning would you say is informal? What do you see as the driving factors that give rise to informal learning?

8. How can plasticity be better supported in the teams, hospitals, and organizations you work in?
